# Depression Risk With PCSK9 Inhibitors Versus Statins in Hyperlipidemia

**DOI:** 10.64898/2026.04.05.26350195

**Authors:** Min-Jing Lee, Chia-Jung Li, Renin Chang, Yen-Feng Lin, Chien-Wei Huang

**Author notes:** **Correspondence author: Yen-Feng Lin, MD, PhD and Chien-Wei Huang, MD,** Yen-Feng Lin, MD, PhD, Center for Neuropsychiatric Research, National Health Research Institutes, Miaoli, Taiwan, No.35, Keyan Road, Zhunan Town, Miaoli County 35053, Taiwan, Chien-Wei Huang, MD, Division of Nephrology, Department of Internal Medicine, Kaohsiung Veterans General Hospital, Kaohsiung, Taiwan, No.386, Dazhong 1st Rd., Zuoying Dist., Kaohsiung City 813414, Taiwan.

## Abstract

**Background:** Hyperlipidemia is a major risk factor for cardiovascular disease and is increasingly linked to depression, which is associated with adverse cardiovascular prognosis. As proprotein convertase subtilisin/kexin type 9 (PCSK9) inhibitors are increasingly used for lipid lowering, their neuropsychiatric safety profile compared with established therapies remains uncertain.

**Objectives:** This study aimed to compare the risk of incident depression associated with initiation of PCSK9 inhibitor therapy vs statin therapy among adults with hyperlipidemia.

**Methods:** In this population-based cohort study, we emulated a target trial using a new-user active-comparator design and real-world data from the TriNetX research network from July 1, 2020, to June 30, 2025. Adults with hyperlipidemia who newly initiated PCSK9 inhibitors or statins were included. The exposure was initiation of PCSK9 inhibitor therapy versus statin therapy. Propensity score matching was performed, yielding 17,805 patients in each group. The primary outcome was incident depression. Cumulative incidence was estimated using the Kaplan-Meier method, and hazard ratios (HRs) with 95% confidence intervals (Cis) were estimated using Cox proportional hazards models.

**Results:** Among 35 610 propensity score–matched patients, the mean age was 65.4 (10.6) years and 46.7% were female. During a mean follow-up of 35.0 (21.2) months, incident depression occurred in 546 patients (3.1%) initiating PCSK9 inhibitors and 981 patients (5.5%) initiating statins. The 5-year cumulative incidence of depression was 5.84% for PCSK9 inhibitor initiators and 7.91% for statin initiators. PCSK9 inhibitor initiation was associated with a lower risk of incident depression (HR, 0.74; 95% CI, 0.67–0.82), corresponding to a 5-year number needed to treat of 46. The association was observed for major depressive disorder (HR, 0.71; 95% CI, 0.63–0.80) but not for dysthymic disorder or adjustment disorder. Consistent associations were observed across prespecified subgroups and sensitivity analyses, and the lower depression risk associated with PCSK9 inhibitor initiation remained regardless of comparator statin intensity or lipophilicity.

**Conclusions:** In this real-world target trial emulation, initiation of PCSK9 inhibitor therapy was associated with a lower risk of incident depression compared with statin therapy among adults with hyperlipidemia. Further prospective studies are warranted to confirm these findings and clarify underlying mechanisms.

## Introduction

The global burden of dyslipidemias has increased over the past 30 years.^1^ Elevated plasma low density lipoprotein cholesterol (LDL-C) levels is a well-established risk factor for atherosclerotic cardiovascular disease, a leading cause of morbidity and mortality worldwide.^2^ In addition, increasing attention has been given to the association between hyperlipidemia, cardiovascular diseases and depression. A growing body of evidence suggests that patients with hyperlipidemia and cardiovascular disease exhibited a higher risk of depression.^3–8^ Depression is not merely a comorbid condition but acts as an independent risk factor for adverse cardiovascular outcomes, significantly impairing long-term survival in patients with established heart disease.^3^ This underscores a complex, bidirectional relationship between metabolic dysregulation and mental health, mediated by shared pathways such as chronic inflammation, hypothalamic-pituitary-adrenal axis dysregulation, altered monoaminergic metabolism, changes in serotonin receptor dynamics, and autonomic imbalance.^7–10^

Pharmacologic regulation of LDL-C is a cornerstone of therapy to reduce cardiovascular events and premature mortality. Hydroxymethylglutaryl Coenzyme A (HMG-CoA) reductase inhibitors, commonly known as statins, have been the mainstay of hyperlipidemia treatment for many years.^11^ Evidence indicates that statins influence neurobiological, cardiometabolic, and immunological pathways implicated in depression.^12,13^ Prior research have suggested potential antidepressant effects of statins, largely attributed to their anti-inflammatory properties.^13–15^ In contrast, some studies have proposed that low cholesterol may contribute to a decrease in brain serotonin, which is associated with depressive symptoms.^5,12,16–18^ Clinical studies investigated on the relationship between statin use and depression revealed inconsistent finding.^19–23^ For example, one Mendelian randomization study suggested an increased risk of depression associated with both statins and proprotein convertase subtilisin/kexin type 9 (PCSK9) inhibitors,^20^ whereas subsequent meta-analyses found no association or even a decreased risk with statin use.^22,23^

Over the past decade, PCSK9 inhibitors have been documented to be the most potent lipid-lowering agents and the prescriptions in the United States increased 2.7-fold from 2015 to 2021.^24–26^ In addition to regulation of cholesterol metabolism through degradation of hepatic LDL receptors, diverse function of PCSK9 in the kidney, pancreas, intestine, and central nervous system have been investigated.^27,28^ Emerging evidence suggests that PCSK9 play a role in neurogenesis, neural cell differentiation, central LDL receptor metabolism, neural cell apoptosis and neuroinflammation.^29,30^ The safety concerns attributed to their effect of lowering LDL cholesterol level has been linked to several neuropsychiatric disorders such as Alzheimer’s disease, alcohol use disorder, stroke, and depression.^31^ Studies examining the effect of PCSK9 on depression, however, have produced conflicting results.^20,32–35^ Data from real world environment to explore the relationship between PCSK9 inhibitor use and depression is still lacking.

As statins and PCSK9 inhibitors represent as the established primary therapy and as an emerging treatment for hyperlipidemia, it is clinically relevant and warranted to directly compare their respective risks of depression. While low cholesterol levels have historically been linked to depression, the distinct mechanism of PCSK9 inhibition—particularly its effects on neuroinflammatory pathways—raises the possibility of a different neuropsychiatric profile compared with statins. To address this knowledge gap, we conducted a target trial emulation with a new-user active comparator design using real-world and population-based data to examine the longitudinal association between PCSK9 inhibitor use and the risk of depression, in comparison with statin use.

## Methods

### Data Source

This study was conducted using a real-world research database from the TriNetX US Collaborative Network. This network provides deidentified electronic health records (EHRs) from academic medical centers and their affiliated hospitals and outpatient clinics, encompassing data from over 100 million patients across 72 healthcare organizations. This database is integrated into the US Food and Drug Administration’s Real-World Evidence Data Enterprise for drug safety surveillance.^36^

The available data include patient demographics, medical utilization records, diagnoses [coded based on International Classification of Diseases, 10^th^ Revision Clinical Modification codes (ICD-10-CM)], medications (coded with Anatomical Therapeutic Chemical Classification System, Veterans Affairs Drug Classification System, or RxNorm), procedures (categorized with International Classification of Diseases, 10^th^ Revision Procedure Coding System or Current Procedural Terminology), and laboratory tests (organized according to Logical Observation Identifiers Names and Codes) with the corresponding test results. Patient-level data analyses were analyzed within the platform using a pre-specified protocol, and the deidentified results were provided to the investigators in a summarized format.^37^

Ethical approval for data analysis was granted from the Institutional Review Board of Kaohsiung Veterans General Hospital (KSVGH24-CT3-09). This study adhered to the ethical principles of the Declaration of Helsinki and reported according to the Strengthening the Reporting of Observational Studies in Epidemiology (STROBE) guideline, and the Transparent Reporting of Observational Studies Emulating a Target Trial (TARGET) guideline.^38^

### Study Design, Eligibility Criteria, and Treatment Strategies

We implemented a target trial emulation using a new-user active comparator design to strengthen causal interpretations from observational data, mirroring a randomized controlled trial. (Supplemental Table 1). We compared the risk of incident depression among patients with hyperlipidemia initiating PCSK9 inhibitors vs statins. Individuals with at least one diagnosis of hyperlipidemia (ICD-10-CM code E78) between July 1, 2020, and June 30, 2025 were enrolled. Patients were classified into two groups based on their prescriptions for hyperlipidemia. The PCSK9 inhibitor group included those who initiated PCSK9 inhibitors following the diagnosis of hyperlipidemia, whereas the statin group included those prescribed statins after the diagnosis of hyperlipidemia. The index date was defined as the date of the first prescription for either PCSK9 inhibitors or statins. Patients younger than 18 years were excluded. We also excluded those with a history of mood disorders or any antidepressant prescription on or before the index date to avoid misclassifying incident depression. Additional exclusions included patients with major psychiatric conditions (schizophrenia, schizoaffective disorder, or anxiety), dementia, neoplasm, or a recorded death on or before the index date. To ensure a new-user active comparator design, the PCSK9i group excluded patients with prior use of statins on or before the index date and PCSK9i before the index date. Conversely, the statin group excluded those with prior use of PCSK9i on or before the index date and statins before the index date. Follow-up began on the index date and continued until the occurrence of the outcome, the final recorded visit, or up to 5 years, whichever occurred first (Supplemental Figure 1). The specific codes used to define the cohorts are provided in Supplemental Table 2.

### Covariates

To balance baseline characteristics, we included covariates assessed within one year before the index date. These comprised demographic variables (age, sex, race), socioeconomic status, lifestyle factors, and medical utilization. Socioeconomic status and lifestyle factors were identified using specific codes. Clinical covariates included comorbidities such as hypertension, ischemic heart disease (IHD), cerebrovascular disease (CVA), overweight and obesity, type 2 diabetes mellitus (T2DM), and chronic kidney disease (CKD). Key medications were also adjusted for, including benzodiazepines, fibrates, ezetimibe, aspirin, angiotensin-converting enzyme inhibitors (ACEi) or angiotensin receptor blockers (ARB), beta blockers, nonsteroidal anti-inflammatory drugs (NSAID), sodium-glucose cotransporter-2 (SGLT2) inhibitors, and glucagon-like peptide-1 receptor agonists (GLP-1 RA). Laboratory data from TriNetX represented the most recent measurements obtained prior to the index date. We included body weight index (BMI, ≥30 kg/m2), and lipid measures [total cholesterol (≥200 mg/dL), LDL-C (≥130 mg/dL), high-density cholesterol (HDL-C, <40 mg/dL), and triglyceride (≥200 mg/dL)]. Detailed covariate coding is provided in Supplemental Table 3.

### Prespecified Outcomes

The primary outcome was the incidence of depression, identified using ICD-10-CM codes (F32.0-32.3, F32.8-32.9, F33.0-33.3, F34.1 and F43.21). All-cause mortality was defined by the comprehensive TriNetX “Deceased” term, which incorporates hospital death records (including ICD-10-CM code R99) and linkage to external mortality sources. We further assessed the incidence of specific depression subtypes, including major depressive disorder, dysthymic disorder and adjustment disorder. To validate our approach, we conducted a positive control outcome analysis using T2DM as a known adverse risk of statins. Furthermore, we performed negative control outcome analyses using outcomes with no expected association with either medication, including acute appendicitis, melanoma and epistaxis, to examine the specificity of the findings. A detailed list of all outcome codes is provided in Supplemental Table 4.

### Statistical Analysis

Baseline characteristics were reported as means with standard deviations (SD) or as counts with percentages. We performed 1:1 propensity score matching (PSM) using the greedy nearest neighbor method with a caliper width of 0.1, and covariate balance was confirmed when standardized mean differences (SMD) were < 0.10. To address the missing data in baseline laboratory variables, a dedicated “missingness” category was created for each variable and incorporated into the PSM model. Outcome incidence was estimated using the Kaplan-Meier method, with differences assessed by the log-rank test. The 5-year cumulative incidence was estimated using the Kaplan–Meier method with right censoring. The number needed to treat (NNT) was calculated as the inverse of the absolute risk difference in cumulative incidence between the two cohorts. Hazard ratios (HRs) and 95% confidence intervals (CIs) were obtained from Cox proportional hazards model. The proportional hazards assumptions were evaluated using the generalized Schoenfeld method; if the assumption was violated, HRs were obtained from a time-varying Cox model. The potential impact of unmeasured confounding was quantified using the E-value.^39^ Duration-specific sensitivity analyses with follow-up truncated at 0.5, 1, and 3 years were further performed to ensure robustness of our findings across different observation periods.

Prespecified subgroup analyses were performed according to age (<65 or ≥65 years), sex (male or female), hypertension, CVA, T2DM, and use of ACEi/ARB, SGLT2 inhibitors, and GLP-1 RA. Furthermore, to assess the differential effects of the active comparator, we conducted separate subgroup analyses based on statin characteristics. First, statin intensity was categorized as low-, moderate-, or high-intensity based on the expected average reduction in LDL cholesterol (<30% for low-intensity, 30%–49% for moderate-intensity, and ≥50% for high-intensity). Second, statin lipophilicity was classified as either lipophilic statin (atorvastatin, simvastatin, lovastatin, and fluvastatin) or hydrophilic statin (rosuvastatin, pravastatin, and pitavastatin). Interaction tests were used to assess differences in treatment effects across subgroups. Sensitivity analyses were conducted to evaluate the robustness of the primary findings. To address potential protopathic bias, HR was re-estimated using a landmark approach by applying varying grace periods of 3, 6, 9, or 12 months calculated from the index date, with the start of follow-up designated to begin after this period. Cox proportional hazards models were repeated using varied enrollment start dates. We performed Cox proportional hazards analyses using three nested models to systematically assess covariate confounding. Model 1 adjusted for age, sex, and race; model 2 further adjusted for socioeconomic status, lifestyle factors, and healthcare utilization; and model 3 additionally adjusted for comorbidities and medications. We further restricted the statin group to patients prescribed high-intensity statins plus ezetimibe, where ezetimibe use was recorded within one month before or on the index date. We also restricted the PCSK9 inhibitor group to patients treated with PCSK9 inhibitors plus statins, where statin use was recorded within one month before or on the index date. For validation, positive control outcome and negative control outcome analyses were conducted. Data collection and analysis were conducted in November 2025. All tests were 2-sided with *P* < .05. Analyses were conducted on the TriNetX platform and using R, version 4.3.3.

## Results

### Demographic Characteristics

A total of 17 815 patients who newly initiated PCSK9 inhibitors and 3 286 430 patients who newly initiated statins were included (Figure 1). Before PSM, the PCSK9 inhibitor group was older (mean age 65.5 years vs 62.0 years) and showed significant differences in demographic, clinical, and laboratory characteristics compared with the statin group. Specifically, the PCSK9 inhibitor group included a higher proportion of White individuals, a lower proportion of Black or African American individuals, and a lower proportion of Asian individuals. The group also demonstrated greater healthcare utilization of outpatient services but less utilization of emergency department service and preventive medicine services. They also exhibited a higher prevalence of major cardiovascular comorbidities including hypertension, IHD, and CVA, as well as higher use of medications including benzodiazepines, fibrates, ezetimibe, ACEi/ARB, beta-blockers, NSAID, SGLT2 inhibitors and GLP-1 RAs. Additionally, the PCSK9 inhibitor group had a higher proportion of individuals with BMI ≥30 kg/m^2^ and unfavorable lipid profiles, characterized by elevated total cholesterol, LDL-C, and triglycerides. After PSM, each cohort contained 17 805 patients, and all baseline characteristics were balanced, with SMD of <0.1 (Table 1).

**Figure 1.**
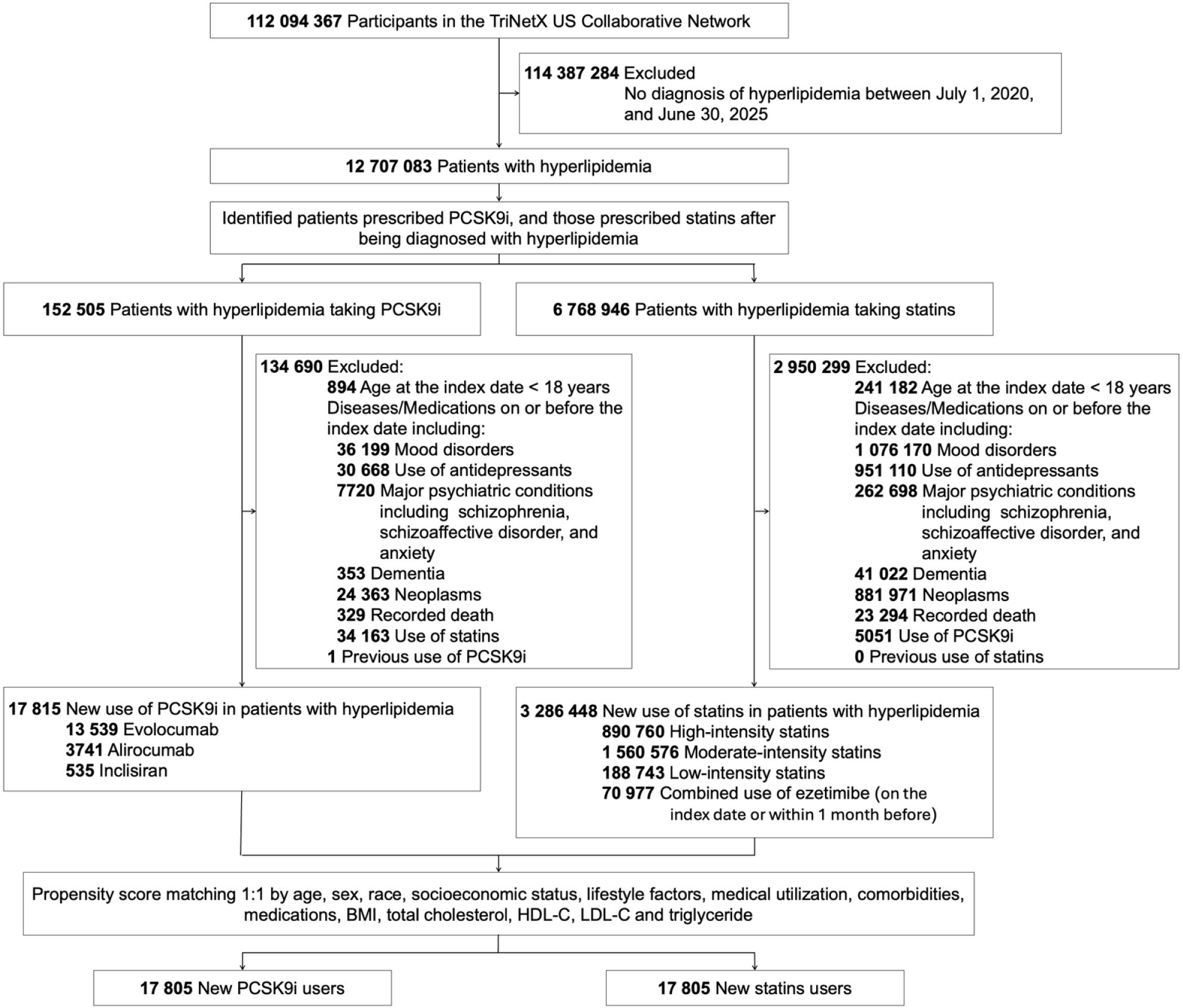
Patient enrollment algorithm. Abbreviations: BMI, body mass index; HDL-C, high-density lipoprotein cholesterol; LDL-C, low-density lipoprotein cholesterol; PCSK9i, proprotein convertase subtilisin/kexin type 9 inhibitors.

**Table 1.** Baseline characteristics of patients with hyperlipidemia treated with PCSK9 inhibitors vs statins.

| Characteristic | Before matching, No. (%) |  |  | After matching, No. (%) |  |  |
| --- | --- | --- | --- | --- | --- | --- |
|  | PCSK9i<br>(n = 17 815) | Statins<br>(n = 3 286 430) | SMD | PCSK9i<br>(n = 17 805) | Statins<br>(n = 17 805) | SMD |
| **Demographic** |  |  |  |  |  |  |
| Age at index, mean (SD), y | 65.5 (10.4) | 62.0 (12.0) | **0.305** | 65.4 (10.4) | 65.4 (10.8) | 0.007 |
| Female, n (%) | 8325 (46.7) | 1 317 497 (40.5) | **0.127** | 8317 (46.7) | 8306 (46.7) | 0.001 |
| **Race, n (%)** |  |  |  |  |  |  |
| White | 13 888 (78.0) | 2 194 162 (67.4) | **0.239** | 13 880 (78.0) | 13 873 (77.9) | 0.001 |
| Black or African American | 1415 (7.9) | 475 260 (14.6) | **0.212** | 1415 (7.9) | 1662 (9.3) | 0.049 |
| Asian | 573 (3.2) | 195 172 (6.0) | **0.133** | 573 (3.2) | 708 (4.0) | 0.041 |
| **Socioeconomic status, n (%)** |  |  |  |  |  |  |
| Persons with potential health hazards related to socioeconomic and psychosocial circumstances | 43 (0.2) | 16 698 (0.5) | 0.044 | 43 (0.2) | 39 (0.2) | 0.005 |
| Problems related to education and literacy | 10<sup>a</sup> (0.1) | 943 (<0.1) | 0.013 | 10<sup>a</sup> (0.1) | 10<sup>a</sup> (0.1) | < .001 |
| Problems related to employment and unemployment | 10<sup>a</sup> (0.1) | 1404 (<0.1) | 0.006 | 10<sup>a</sup> (<0.1) | 10<sup>a</sup> (<0.1) | < .001 |
| Problems related to housing and economic circumstances | 12 (0.1) | 8008 (0.2) | 0.045 | 12 (0.1) | 12 (0.1) | < .001 |
| Other problems related to primary support group, including family circumstances | 13 (0.1) | 2941 (0.1) | 0.006 | 13 (0.1) | 10<sup>a</sup> (0.1) | 0.007 |
| **Lifestyle factors, n (%)** |  |  |  |  |  |  |
| Nicotine dependence | 559 (3.1) | 148 094 (4.5) | 0.073 | 559 (3.1) | 548 (3.1) | 0.004 |
| Alcohol related disorders | 74 (0.4) | 30 485 (0.9) | 0.064 | 74 (0.4) | 74 (0.4) | < .001 |
| **Healthcare utilization, n (%)** |  |  |  |  |  |  |
| Office or other outpatient services | 5126 (28.8) | 770 314 (23.7) | **0.117** | 5121 (28.8) | 4743 (26.6) | 0.048 |
| Emergency department services | 1030 (5.8) | 304 782 (9.4) | **0.136** | 1030 (5.8) | 880 (4.9) | 0.037 |
| Hospital inpatient and observation care service | 696 (3.9) | 149 367 (4.6) | 0.034 | 696 (3.9) | 602 (3.4) | 0.028 |
| Preventive medicine services | 574 (3.2) | 209 182 (6.4) | **0.150** | 573 (3.2) | 907 (5.1) | 0.094 |
| **Comorbidities, n (%)** |  |  |  |  |  |  |
| Hypertension | 6582 (36.9) | 996 375 (30.6) | **0.135** | 6576 (36.9) | 6253 (35.1) | 0.038 |
| Ischemic heart disease | 5662 (31.8) | 337 198 (10.4) | **0.545** | 5653 (31.8) | 5759 (32.3) | 0.013 |
| Cerebrovascular disease | 1233 (6.9) | 135 168 (4.2) | **0.121** | 1230 (6.9) | 1226 (6.9) | 0.001 |
| Overweight and obesity | 1445 (8.1) | 274 394 (8.4) | 0.011 | 1443 (8.1) | 1261 (7.1) | 0.039 |
| Type 2 diabetes mellitus | 2694 (15.1) | 518 966 (15.9) | 0.023 | 2691 (15.1) | 2472 (13.9) | 0.035 |
| Chronic kidney disease | 886 (5.0) | 146 526 (4.5) | 0.022 | 886 (5.0) | 812 (4.6) | 0.020 |
| **Medications, n (%)** |  |  |  |  |  |  |
| Benzodiazepine, anxiolytics | 579 (3.3) | 80 221 (2.5) | 0.047 | 579 (3.3) | 563 (3.2) | 0.005 |
| Benzodiazepine, hypnotics and sedatives | 1257 (7.1) | 137 955 (4.2) | **0.122** | 1256 (7.1) | 1142 (6.4) | 0.026 |
| Fibrates | 425 (2.4) | 29 035 (0.9) | **0.118** | 425 (2.4) | 401 (2.3) | 0.009 |
| Ezetimibe | 2322 (13.0) | 19224 (0.6) | **0.510** | 2313 (13.0) | 2279 (12.8) | 0.006 |
| Aspirin | 1992 (11.2) | 203 709 (6.3) | **0.175** | 1990 (11.2) | 1787 (10.0) | 0.037 |
| ACEi/ARB | 3788 (21.3) | 504 390 (15.5) | **0.150** | 3782 (21.2) | 3649 (20.5) | 0.018 |
| β-blockers | 3678 (20.6) | 376 308 (11.6) | **0.249** | 3672 (20.6) | 3659 (20.6) | 0.002 |
| NSAID | 1675 (9.4) | 301 019 (9.2) | **0.006** | 1674 (9.4) | 1561 (8.8) | 0.022 |
| SGLT2 inhibitors | 578 (3.2) | 42 876 (1.3) | **0.129** | 575 (3.2) | 556 (3.1) | 0.006 |
| GLP-1 RA | 644 (3.6) | 49 721 (1.5) | **0.132** | 640 (3.6) | 614 (3.5) | 0.008 |
| **Laboratory tests and examinations** |  |  |  |  |  |  |
| Body mass index ≥30 kg/m<sup>2</sup> | 4172 (23.4) | 622 345 (19.1) | **0.106** | 4168 (23.4) | 3961 (22.2) | 0.028 |
| Missing, n (%) | 9580 (53.8) | 2076523 (63.2) | **0.192** | 9578 (53.8) | 10200 (57.3) | 0.070 |
| Total cholesterol ≥200 mg/dL | 3640 (20.4) | 518 333 (15.9) | **0.117** | 3633 (20.4) | 3107 (17.5) | 0.076 |
| Missing, n (%) | 12 125 (68.1) | 2 311 007 (70.3) | 0.049 | 12 122 (68.1) | 12 017 (67.5) | 0.012 |
| LDL-C ≥130 mg/dL | 3311 (18.6) | 438 444 (13.5) | **0.140** | 3304 (18.6) | 2847 (16.0) | 0.068 |
| Missing, n (%) | 12 017 (67.5) | 2 316 467 (70.5) | 0.066 | 12 014 (67.5) | 11 954 (67.1) | 0.008 |
| HDL-C <40 mg/dL | 1602 (9.0) | 289 859 (8.9) | 0.003 | 1599 (9.0) | 1397 (7.8) | 0.041 |
| Missing, n (%) | 11 914 (66.9) | 2 297 230 (69.9) | 0.065 | 11 911 (66.9) | 11 929 (67.0) | 0.002 |
| Triglyceride ≥200 mg/dL | 1820 (10.2) | 237 570 (7.3) | **0.104** | 1814 (10.2) | 1597 (9.0) | 0.041 |
| Missing, n (%) | 11 011 (61.8) | 2 304 411 (70.1) | **0.176** | 11 908 (66.9) | 11 963 (67.2) | 0.007 |

<sup>a</sup> For confidentiality, TriNetX replaces single-digit counts with 10. **Abbreviations:** ACEi,
angiotensin-converting enzyme inhibitors; ARB, angiotensin receptor blockers; CCB, calcium
channel blockers; GLP-1 RA, glucagon-like peptide-1 receptor agonists; HDL-C, high-density
lipoprotein cholesterol; LDL-C, low-density lipoprotein cholesterol; NSAID, nonsteroidal
anti-inflammatory drugs; PCSK9i, proprotein convertase subtilisin/kexin type 9 inhibitors; SGLT2,
sodium-glucose cotransporter-2; SMD, standardized mean difference.

### Incidence and Risk of Depression

During a mean (±SD) follow-up of 35.0 (±21.2) months, depression developed in 546 patients (3.1%) in the PCSK9 inhibitor group and 981 patients (5.5%) in the statin group (Table 2). Over 5 years, the cumulative incidence of depression was significantly lower in the PCSK9 inhibitor group (5.84%; 95% CI, 5.32%−6.41%) than in the statin cohort (7.91%; 95% CI, 7.43%−8.42%; *P* < .001; Figure 2), corresponding to a NNT of 46. In Cox regression analyses, PCSK9 inhibitor use was associated with a lower risk of depression (HR, 0.74; 95% CI, 0.67−0.82; Table 2). The E-value for the primary outcome was 2.04 (1.74 for the upper bound of the 95% CI). Duration-specific sensitivity analyses revealed that the finding of lower risk depression in the PCSK9 inhibitor group was evident at the 180-day follow-up (HR, 0.60; 95% CI, 0.47–0.75), and this protective association remained consistent across all longer follow-up durations (Table 2). There was no difference in the risk of all-cause mortality between the two groups (HR, 0.92; 95% CI, 0.81−1.04). Results for specific depression subtypes showed that the 5-year cumulative incidence of major depressive disorder was significantly lower in the PCSK9 inhibitor group (4.42%; 95% CI, 3.98%−4.91%) than in the statin cohort (6.44%; 95% CI, 6.00%−6.90%; *P* < .001), corresponding to a NNT of 50. PCSK9 inhibitor use was significantly associated with a lower risk of major depressive disorder compared with statin use (HR, 0.71; 95% CI, 0.63−0.80; Table 2). However, no significant differences were observed in the risks of dysthymic disorder (HR, 0.67; 95% CI, 0.45−1.00) or adjustment disorder (HR, 0.91; 95% CI, 0.70−1.17) between the two groups.

**Figure 2.**
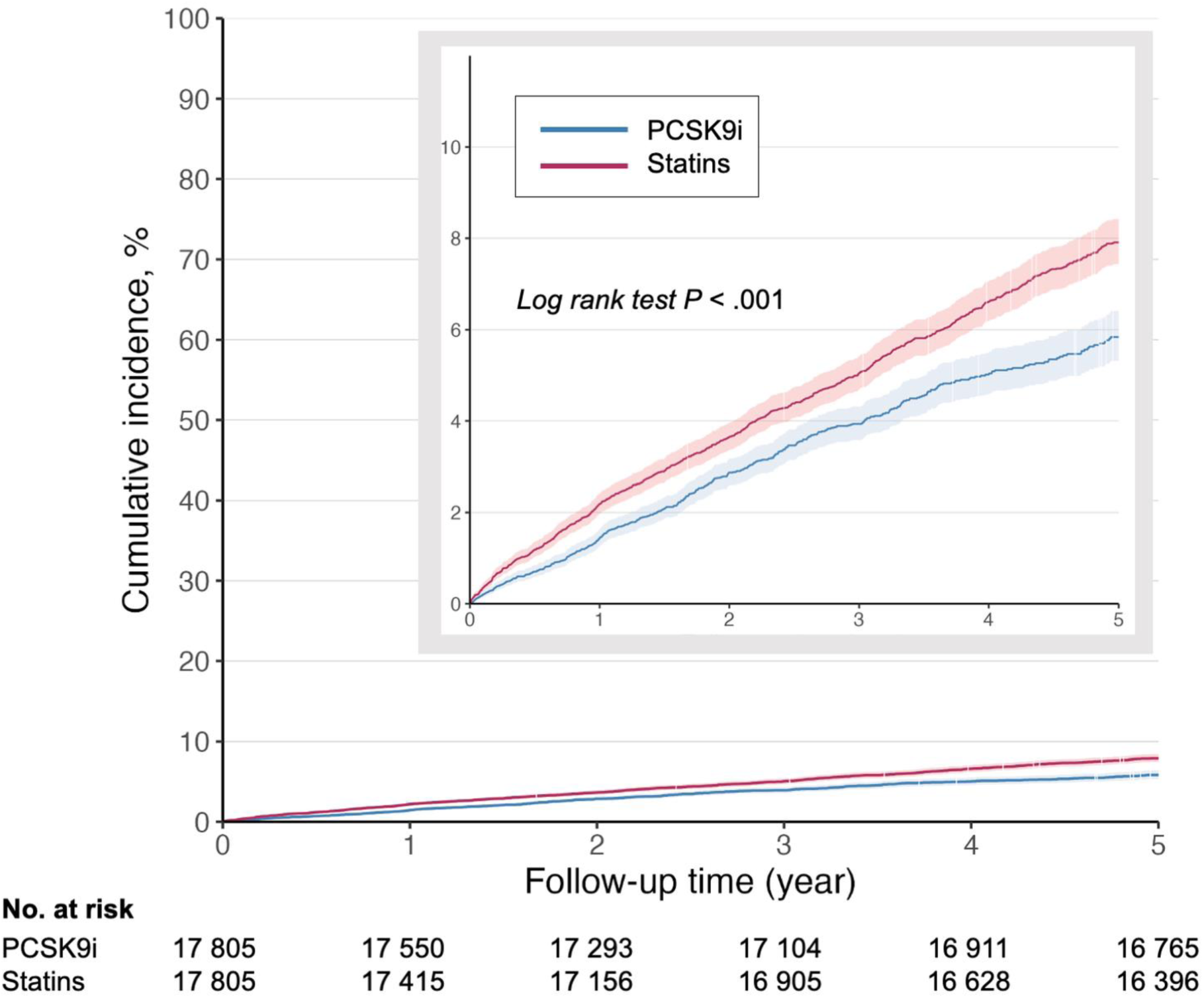
Cumulative incidence of depression in patients with hyperlipidemia treated with PCSK9 inhibitors vs statins. The 5-year cumulative incidences of depression were 5.84% in the PCSK9 inhibitor group and 7.91% in the statin group. The inset presents the same data on an expanded y axis. **Abbreviations:** PCSK9i, proprotein convertase subtilisin/kexin type 9 inhibitors.

**Table 2.**
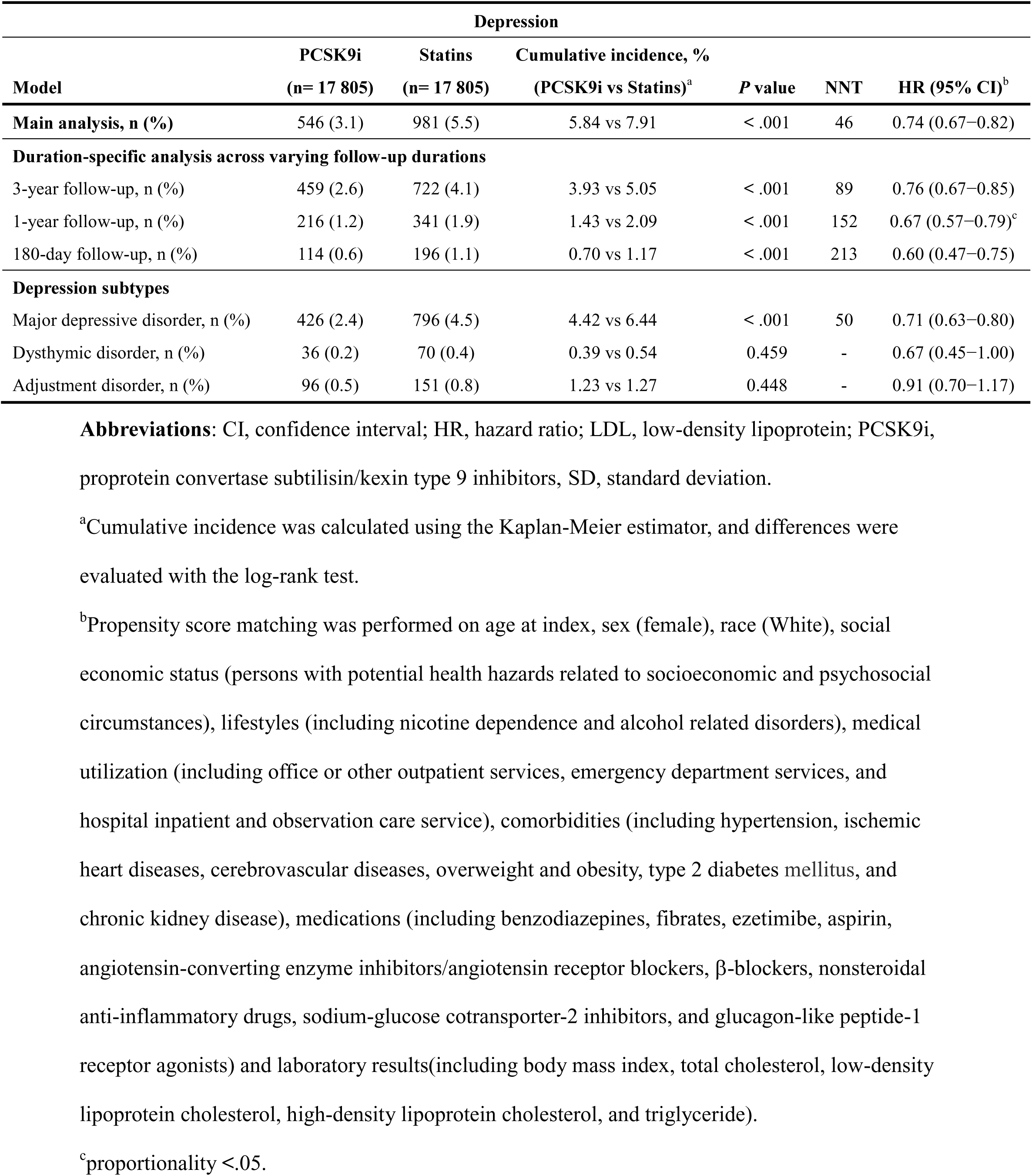
Incidence and risk of depression in patients with hyperlipidemia treated with PCSK9i vs statins.

### Subgroup Analyses

Subgroup analyses stratified by age, sex, hypertension, CVA, T2DM, and medication use including ACEi/ARB, SGLT2 inhibitors, and GLP-1 RA showed that the reduced risk of incident depression in the PCSK9 inhibitor group compared with the statin group was generally consistent across all subgroups (Figure 3). We further assessed the consistency of the lower risk association by stratifying the statin group according to statin dosage intensity and lipophilicity. The reduced risk of depression associated with PCSK9 inhibitors was not significantly modified by either statin intensity (*P* for interaction = .628) or statin lipophilicity (*P* for interaction = .152; Figure 3).

**Figure 3.**
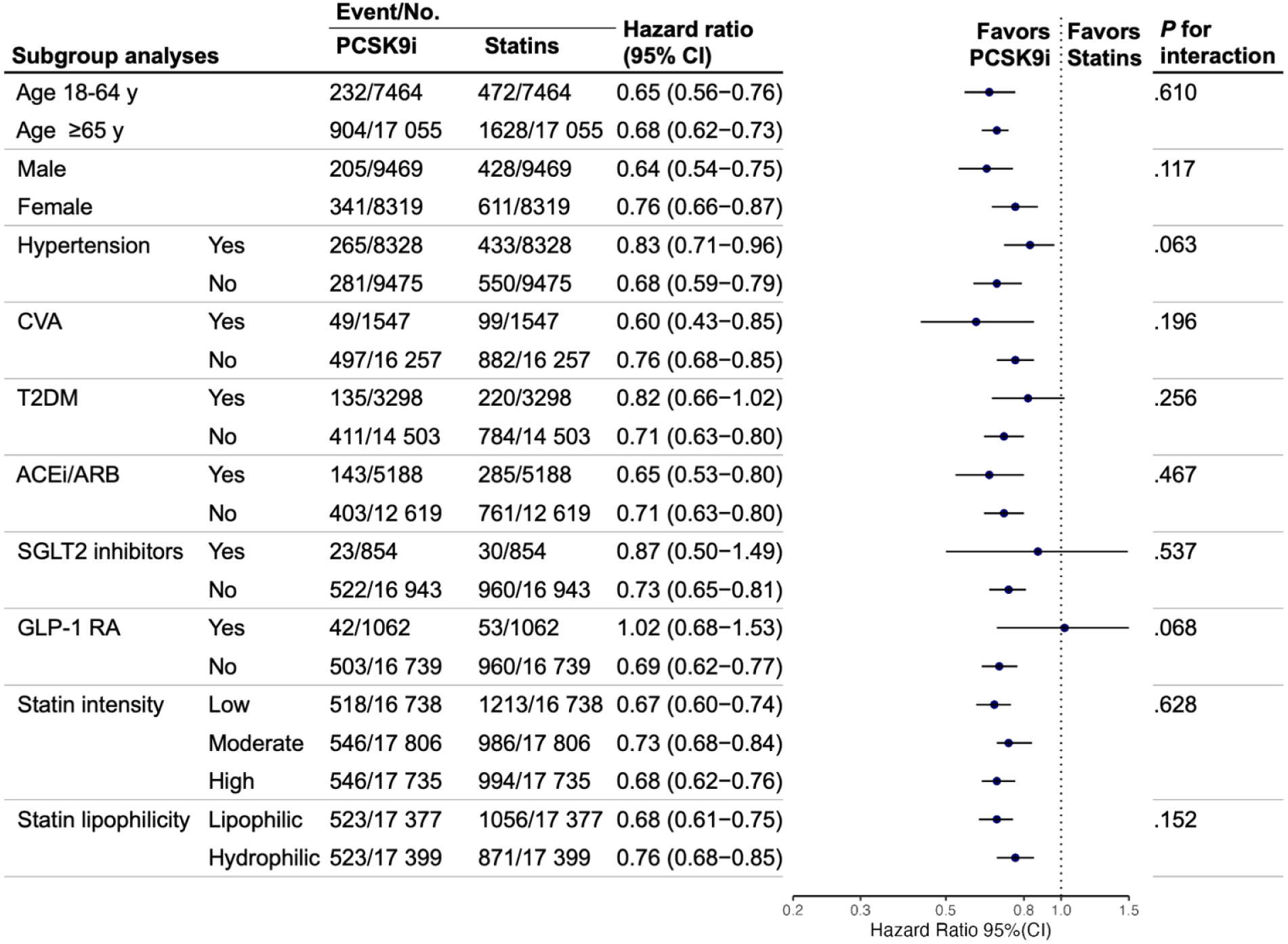
Subgroup analyses of hazard ratios for depression in patients with hyperlipidemia treated with PCSK9 inhibitors vs statins. Statin intensity was categorized based on the expected average reduction in LDL cholesterol: high-intensity statin was defined as ≥ 50% reduction (atorvastatin 40–80 mg, and Rosuvastatin 20–40 mg); moderate-intensity statin was defined as approximately 30%–49% reduction (atorvastatin 10–20 mg, rosuvastatin 5–10 mg, simvastatin 20–40 mg, pravastatin 40-80mg, lovastatin 40mg, fluvastatin 80mg, and pitavastatin 2–4 mg); and low-intensity was defined as < 30% reduction (simvastatin 10 mg, pravastatin 10–20 mg, lovastatin 20mg, and fluvastatin 20–40mg). Statin lipophilicity was classified as lipophilic statin (atorvastatin, simvastatin, lovastatin, and fluvastatin) and hydrophilic statin (rosuvastatin, pravastatin, and pitavastatin). **Abbreviations:** ACEi, angiotensin-converting enzyme inhibitors; ARB, angiotensin receptor blockers; CVA, cerebrovascular disease; GLP-1RA, glucagon-like peptide-1 receptor agonists; PCSK9i, proprotein convertase subtilisin/kexin type 9 inhibitors; SGLT2, sodium-glucose cotransporter-2; T2DM, type 2 diabetes mellitus.

### Sensitivity and Validation Analyses

Sensitivity analyses using the landmark approach by applying different grace periods for the start of the follow-up (3, 6, 9, and 12 months after the index date) and varied enrollment start dates yielded results consistent with the primary analysis (HR ranges: 0.72−0.74 and 0.66−0.73, respectively; Supplemental Figure 2). The primary results remained stable across the three nested Cox models with sequential covariate adjustment (HR ranges: 0.71−0.73; Supplemental Figure 2). Results were further consistent when restricting the cohorts based on the co-medication use of high-intensity statins plus ezetimibe (HR, 0.76) or PCSK9 inhibitors plus statins (HR, 0.54; Supplemental Figure 2). Collectively, these sensitivity analyses provided further support for the primary finding of a lower risk of depression associated with PCSK9 inhibitor use. In the positive control outcome analysis, the risk of incident T2DM was significantly lower in the PCSK9 group compared with the statin group (HR, 0.86; 95% CI, 0.81−0.92). The negative control outcome analyses using acute appendicitis, melanoma and epistaxis showed no significant associations with either group (Supplemental Figure 2).

## Discussion

To our knowledge, this is the first target trial emulation using real-world, population-based data to investigate the risk of depression among patients with hyperlipidemia initiating PCSK9 inhibitors vs statins under a new-user active comparator design. We provide robust evidence that PCSK9 inhibitor therapy is associated with a significantly lower risk of incident depression compared with statin therapy. The decreased depression risk associated with PCSK9 inhibitor use was evident as early as 180 days and persisted across follow-up durations. When examining specific outcomes, a significant risk reduction was observed for major depressive disorder but not for dysthymic or adjustment disorder, a pattern that supports a biologically mediated effect rather than psychosocial influences. Importantly, patients initiating PCSK9 inhibitors in our cohort had a higher baseline burden of cardiovascular comorbidities—conditions that are themselves associated with an increased risk of depression. Despite this unfavorable baseline risk profile, PCSK9 inhibitor use was associated with a lower incidence of depression, which strengthens the credibility of the observed association. Notably, this imbalance would be expected to bias the observed association toward the null rather than generate a spurious protective effect, thereby reinforcing the causal interpretation of our findings. Consistent associations were observed across various subgroups, and this protective association remained regardless of the comparator’s statin intensity or lipophilicity. Sensitivity and validation analyses, including positive and negative control outcome analyses, further confirmed the robustness of these findings. These results suggest that PCSK9 inhibitor therapy may have a more favorable neuropsychiatric safety profile with respect to depression risk compared with statin therapy.

Previous concerns regarding the neuropsychiatric safety of PCSK9 inhibitors stem from the hypothesis that cholesterol reduction could alter neuronal membrane fluidity and serotonin receptor function, potentially contributing to impaired cognitive function and depressive symptoms.^31,40^ Sporadic cases of depression have been reported in pharmacovigilance systems, but large-scale genetic and clinical studies have yielded inconsistent findings.^35,40^ Two Mendelian randomization analyses from the largest genome-wide association studies of European ancestry suggested that genetically reduced PCSK9 expression was not associated with an increased risk of major depression, mood instability, or neuroticism.^32,33^ In contrast, a study by Alghamdi et al., based on Mendelian randomization using data from the Global Lipid Genetics Consortium, reported an increased risk of depression with both statins and PCSK9 inhibitors, with a slightly higher and more significant risk for PCSK9 inhibitors.^20^ Methodological differences may explain the divergence between our findings and these genetic studies. Mendelian randomization analyses use genetic variants as proxies for the lifelong systemic effects of reduced PCSK9 function, whereas current PCSK9 inhibitors act peripherally on circulating LDL receptors and represent a relatively short-term pharmacological intervention. Our findings therefore complement the genetic evidence by providing direct, real-world data on the neuropsychiatric safety profile of PCSK9 inhibitor therapy. Moreover, the observed reduction in depression risk was confined to major depressive disorder, with no effect on dysthymic or adjustment disorders. This differential pattern supports that the association may be biologically mediated rather than driven by psychosocial factors. This divergence offers valuable insight for causal interpretation, emphasizing the need to distinguish biologically mediated effects from residual population-level confounding.

The inconclusive evidence regarding the effect of statins on depression may be explained by heterogeneity across studies, distinct pharmacologic properties such as lipophilicity and blood–brain barrier permeability.^23^ In our study, PCSK9 inhibitor therapy was associated with a lower risk of depression compared with both lipophilic and hydrophilic statins, and this protective association persisted across all levels of statin intensity. Notably, patients receiving combined PCSK9 inhibitor and statin therapy exhibited the lowest risk of depression. This pattern argues against the notion that the observed association is driven solely by the adverse neuropsychiatric effects of lipophilic statins penetrating the blood–brain barrier. Instead, it suggests that PCSK9 inhibitors may possess distinct pharmacologic properties relevant to mood regulation. These properties may include limited central nervous system penetration and systemic anti-inflammatory effects, as well as reductions in lipoprotein(a) concentrations, which together could confer neuroprotective benefits beyond lipid lowering.^29,41^

### Strengths and Limitations

This study has several notable strengths. First, the implementation of a target trial emulation with a new-user active comparator design minimized common biases in observational research, including immortal time and prevalent user bias. This design is particularly important in the context of PCSK9 inhibitors, which were introduced later than statins, as comparisons that include prevalent statin users may otherwise be prone to survivor and selection biases. Second, the use of a large, real-world database enhanced statistical power and generalizability, and inclusion of key laboratory parameters such as BMI and lipid profiles enabled granular PSM to reduce confounding by indication. Third, comprehensive subgroup analyses stratified by statin lipophilicity and intensity provided mechanistic context, allowing partial separation of lipid-lowering effects from potential neurobiological influences. Fourth, the specificity of the association observed for major depressive disorder but not adjustment disorder supports a biological rather than purely psychosocial explanation. Finally, the consistency of findings across extensive sensitivity analyses, coupled with the E-value, NNT assessment, and validation analyses, reinforces the robustness and clinical interpretability of the results. While the E-value suggests that moderate unmeasured confounding could attenuate the observed association, it is unlikely to fully explain the convergence of early temporal separation, outcome specificity for major depressive disorder, and consistently null findings across multiple control outcomes.

Several limitations should be acknowledged. First, reliance on diagnostic codes may have introduced ascertainment bias; however, the results of positive and negative control outcome analyses indicated minimal bias. Moreover, any misclassification of depression diagnoses is likely to be non-differential between treatment groups, which would bias the estimates toward the null rather than account for the observed protective association. Second, drug exposure was based on prescription records rather than actual drug consumption, which prevents direct assessment of patient adherence and treatment persistence. Nevertheless, aligning with the intention-to-treat principle of our target trial emulation, this design robustly estimates the real-world effectiveness of the initial treatment strategy, inherently capturing the impact of non-adherence. Although both cohorts were restricted to new users without prior exposure to the comparator therapy, residual confounding related to health-seeking behavior, treatment adherence, or healthcare utilization—such as a healthy user or adherence effect—cannot be entirely excluded. Finally, validation in additional EHR databases is warranted to confirm the generalizability of these findings.

## Conclusions

In this study using a new-user active comparator design and a target trial emulation framework, PCSK9 inhibitor therapy was associated with a lower risk of developing depression compared with statin therapy among patients with hyperlipidemia. The association was robust across multiple analyses and appeared specific to major depressive disorder, supporting the possibility of distinct neuropsychiatric effects related to PCSK9 inhibition. These observations highlight a potentially important intersection between lipid metabolism and mental health and warrant further prospective and mechanistic studies.

## Abbreviations

HMG-CoA reductase inhibitors: Hydroxymethylglutaryl Coenzyme A reductase inhibitors
LDL-C: low density lipoprotein cholesterol
PCSK9 Inhibitors: proprotein convertase subtilisin/kexin type 9

## Data Availability Statement

All data utilized or analyzed in this study are included in this published article and its supplemental material files. The data supporting the findings of this study were obtained from the TriNetX research network and are subject to third-party restrictions. All data are accessible to accredited researchers through a licensing agreement with TriNetX. Requests for access can be made via the TriNetX platform (https://live.trinetx.com).

## Appendix

For supplemental figures and tables, please see the online version of this paper.

