## Supplemental for "Depression Risk With PCSK9 Inhibitors Versus Statins in Hyperlipidemia"

**Supplemental Table 1.** Specification and emulation of the target trial using real-world data.

**Supplemental Table 2.** Demographic, visit, diagnostic, and medication used in the definition of the cohorts.

**Supplemental Table 3.** Demographic, diagnostic, procedure, medication, and laboratory codes used in the definition of covariates.

**Supplemental Table 4.** Demographics, diagnostic, and laboratory codes used in the definition of outcomes.

**Supplemental Figure 1.** Graphical illustration of the study design.

**Supplemental Figure 2.** Forest plot depicting hazard ratios for depression incidence in sensitivity and validation analyses.

**STROBE Statement**—Checklist of items that should be included in reports of *cohort studies*

**TARGET Checklist of Recommended Items to Address in Reports of Studies Emulating a Target Trial**

Supplemental material has been provided by the authors to offer readers additional insights into their study.

**Supplemental Table 1. Specification and emulation of the target trial using real-world data.**

| **Protocol Component** | **Target Trial** | **Emulated Trial** |
| --- | --- | --- |
| **Aim** | To compare the risk of incident depression among patients with hyperlipidemia initiating PCSK9 inhibitors vs statins. | Same as for the target trial. |
| **Eligibility Criteria** | - Hyperlipidemia - Age ≥ 18 years at cohort entry. - No prior PCSK9 inhibitor or statin usage. - No prior mood disorders - No prior antidepressants usage - No prior major psychiatric conditions (schizophrenia, schizoaffective disorder, and anxiety) - No prior dementia diagnosis - No prior neoplasm diagnosis | Same as for the target trial. |
| **Treatment Strategies** | PCSK9 inhibitor treatment vs statin treatment. | Same as for the target trial. |
| **Treatment Assignment** | Eligible individuals are randomly assigned to the treatment groups (the same probability of treatment assignment between the treatment groups). | Randomization is emulated by 1:1 PSM to generate a study population with similar probability of treatment assignment between groups. |
| **Follow-up** | Follow-up starts at treatment assignment and ends on the day of the first occurrence of the outcome, death, loss of follow-up, a maximum of 5 years, or the end of the study period, whichever occurs first. | - Follow-up starts at treatment initiation and ends on the day of the first occurrence of the outcome, death, loss of follow-up, a maximum of 5 years, or the end of the study period, whichever occurs first (Same as for the target trial). |
| **Outcomes** | - Incident depression - Incident specific depression subtypes (major depressive disorder, dysthymic disorder, adjustment disorder). | Same as for the target trial. |
| **Causal Contrast** | Intention-to-treat effect. | Same (as-started analysis, analog of intention-to-treat). |
| **Identifying assumptions** | The intention-to-treat effect is identifiable under three core assumptions: 1. Consistency: The treatment strategies are sufficiently well-defined. 2. Exchangeability: The randomized assignment ensures baseline exchangeability between the treatment groups. 3. Non-informative Censoring: If there are losses to follow-up, they are non-informative, conditional on baseline variables. | 1. Consistency: treatment of both drug exposure was based on prescriptions written in the electronic health records, and the potential outcome for each participant is unaffected by the treatment received by others. 2. Conditional exchangeability: We assume that the choice of initiating PCSK9 inhibitors vs statins is conditionally independent of outcomes, given the comprehensive set of measured baseline covariates (as specified in Item 7g.ii) used in the propensity score matching. 3. Positivity: For every combination of the measured baseline covariates, there is a non-zero probability of initiating either PCSK9 inhibitors or statins. 4. Non-informative censoring: We assume that administrative censoring and loss to follow-up are independent of the potential outcomes, conditional on the measured covariates. |
|  |  | The baseline variables including demographics (age at index, female, and White), socioeconomic status (persons with potential health hazards related to socioeconomic and psychosocial circumstances), lifestyle factors (nicotine dependence, and alcohol related disorders), medical utilization (office or other outpatient services, emergency department services, and hospital inpatient and observation care service), comorbidities (hypertension, ischemic heart diseases, cerebrovascular diseases, overweight and obesity, type 2 diabetes mellitus, and chronic kidney disease), and medications (benzodiazepines, fibrates, ezetimibe, aspirin, angiotensin-converting enzyme inhibitors/angiotensin receptor blockers, β-blockers, nonsteroidal anti-inflammatory drugs, sodium-glucose cotransporter-2 inhibitors, and glucagon-like peptide-1 receptor agonists) are operationalized using diagnostic, procedure, or drug codes recorded within the 12-month prior to the date of treatment assignment. For laboratory data (body mass index, total cholesterol, low-density lipoprotein cholesterol, high-density lipoprotein cholesterol, and triglyceride), the most recent measurement recorded in the electronic health records before the treatment assignment is obtained, and a missing indicator variable was created for each laboratory variable and included as a categorical covariate in the PSM model to address the missing data. |
| **Data analysis plan** | - The 5-year cumulative incidence of depression and the Number Needed to Treat, derived from the 5-year cumulative risk difference, were calculated for the PCSK9 inhibitor group versus the statin group. - The HR of PCSK9 inhibitors vs statins with its 95% confidence interval were estimated using a Cox proportional hazards model. - It is assumed that the target trial is an ideal randomized trial where the measured baseline covariates are sufficient to control for confounding, and all relevant variables are completely collected without missing data. - The proportional hazards assumption is expected to hold | - The 5-year cumulative incidence of depression and the Number Needed to Treat, derived from the 5-year cumulative risk difference, were calculated for the PCSK9 inhibitor group versus the statin group. (Same as for the target trial). - The HR of PCSK9 inhibitors vs statins with its 95% confidence interval were estimated using a Cox proportional hazards model. (Same as for the target trial). - To address the missing data in baseline laboratory variables, a missing indicator variable was created for each variable and included as a categorical covariate in the PSM model. - The proportional hazards assumption was assessed using Schoenfeld residuals - Additional analyses to assess the robustness of the results:   - Duration-specific sensitivity analyses with follow-up truncated at 0.5, 1, and 3 years   - E-values to evaluate the potential effect of unmeasured confounders   - Re-estimate HR using a landmark approach by applying varying grace periods of 3, 6, 9, or 12 months calculated from the index date, with the start of follow-up designated to begin after this period.   - Repeat Cox proportional hazards models using varied enrollment start dates.   - Perform Cox proportional hazards analyses using three nested models to systematically assess covariate confounding.   - Examine the influence of statin dosage intensity by conducting separate analyses for patients categorized as low-, moderate-, and high-intensity statin users.   - Restrict the statin group to patients prescribed high-intensity statins plus ezetimibe on or one month before the index date.   - Restrict the PCSK9 inhibitor group to patients treated with PCSK9 inhibitors plus statins on or one month before the index date.   - HRs were obtained from a time-varying Cox model when Schoenfeld residuals indicated non-proportional hazards. |

**Abbreviations**: HR, hazard ratio; PCSK9i, proprotein convertase subtilisin/kexin type 9 inhibitors; PSM, propensity score matching.

**Supplemental Table 2.** Demographic, visit, diagnostic, and medication used in the definition of the cohorts.

| **Category** | **Code** | **Description** |
| --- | --- | --- |
| **Hyperlipidemia (specific date: 2020/07/01 - 2025/06/30)** | | |
| Diagnosis | ICD10CM:E78 | Disorders of lipoprotein metabolism and other lipidemias |
| **Patients taking PCSK9 inhibitors or those taking statins after being diagnosed with hyperlipidemia** | | |
| Medication | RxNorm: 1659152 | Alirocumab |
| Medication | RxNorm: 1665684 | Evolocumab |
| Medication | RxNorm: 2588243 | Inclisiran |
| Medication | ATC: C10AA | HMG CoA reductase inhibitors |
| **Exclude age at index date < 18 years** | | |
| Term filters | Age at Event | 18 years or older |
| **Exclude mood disorders on or before the index date** | | |
| Diagnosis | ICD10CM:F30-39 | Mood [affective] disorders |
| **Exclude use of antidepressants on or before the index date** | | |
| Medication | ATC: N06A | Antidepressants |
| **Exclude major psychiatric conditions on or before the index date** | | |
| Diagnosis | ICD10CM:F20 | Schizophrenia |
| Diagnosis | ICD10CM:F25 | Schizoaffective disorders |
| Diagnosis | ICD10CM:F41 | Other anxiety disorders |
| **Exclude dementia on or before the index date** | | |
| Diagnosis | ICD10CM:G30 | Alzheimer’s disease |
| Diagnosis | ICD10CM:F01 | Vascular dementia |
| Diagnosis | ICD10CM:F03 | Unspecified dementia |
| **Exclude neoplasms on or before the index date** | | |
| Diagnosis | ICD10CM:C00-D49 | Neoplasms |
| **Exclude recorded death on or before the index date** | | |
| Demographics | Deceased | Deceased |
| Diagnosis | ICD10CM:R99 | Ill-defined and unknown cause of mortality |
| **Exclude statins on or before the index date or previous use of PCSK9 inhibitors in the PCSK9 inhibitor cohort** | | |
| Medication | ATC: C10AA | HMG CoA reductase inhibitors |
| Medication | RxNorm: 1659152 | Alirocumab |
| Medication | RxNorm: 1665684 | Evolocumab |
| Medication | RxNorm: 2588243 | Inclisiran |
| **Exclude PCSK9 inhibitors on or before the index date or previous use of statins in the statin cohort** | | |
| Medication | RxNorm: 1659152 | Alirocumab |
| Medication | RxNorm: 1665684 | Evolocumab |
| Medication | RxNorm: 2588243 | Inclisiran |
| Medication | ATC: C10AA | HMG CoA reductase inhibitors |

**Abbreviations:** HMG CoA, hydroxymethylglutaryl-coenzyme A; PCSK9, proprotein convertase subtilisin/kexin type 9.

**Supplemental Table 3.** Demographic, diagnostic, procedure, medication, and laboratory codes used in the definition of covariates.

| **Covariates** | **Category** | **Code** | **Description** |
| --- | --- | --- | --- |
| Age at index | Demographics | AI | Age at index |
| Male | Demographics | M | Male |
| Female | Demographics | F | Female |
| White | Demographics | 2106-3 | White |
| Black or African American | Demographics | 2054-5 | Black or African American |
| Asian | Demographics | 2028-9 | Asian |
| Persons with potential health hazards related to socioeconomic and psychosocial circumstances | Diagnosis | ICD10CM:Z55-Z65 | Persons with potential health hazards related to socioeconomic and psychosocial circumstances |
| Problems related to education and literacy | Diagnosis | ICD10CM:Z55 | Problems related to education and literacy |
| Problems related to employment and unemployment | Diagnosis | ICD10CM:Z56 | Problems related to employment and unemployment |
| Problems related to housing and economic circumstances | Diagnosis | ICD10CM:Z59 | Problems related to housing and economic circumstances |
| Other problems related to primary support group, including family circumstances | Diagnosis | ICD10CM:Z63 | Other problems related to primary support group, including family circumstances |
| Nicotine dependence | Diagnosis | ICD10CM:F17 | Nicotine dependence |
| Alcohol related disorders | Diagnosis | ICD10CM:F10 | Alcohol related disorders |
| Office or other outpatient services | Procedures | CPT:1013626 | Office or other outpatient services |
| Emergency department services | Procedures | CPT:1013711 | Emergency department services |
| Hospital inpatient and observation care service | Procedures | CPT:1013659 | Hospital inpatient and observation care service |
| Preventive medicine services | Procedures | CPT:1013829 | Preventive medicine services |
| Hypertension | Diagnosis | ICD10CM:I10 | Essential (primary) hypertension |
| Ischemic heart disease | Diagnosis | ICD10CM:I20-I25 | Ischemic heart disease |
| Cerebrovascular disease | Diagnosis | ICD10CM:I60-I69 | Cerebrovascular diseases |
| Overweight and obesity | Diagnosis | ICD10CM:E66 | Overweight and obesity |
| Type 2 diabetes mellitus | Diagnosis | ICD10CM:E11 | Type 2 diabetes mellitus |
| Chronic kidney disease | Diagnosis | ICD10CM:N18 | Chronic kidney disease (CKD) |
| Benzodiazepine, anxiolytics | Medication | ATC: N05BA | Benzodiazepine, derivatives |
| Benzodiazepine, hypnotics and sedatives | Medication | ATC: N05CD | Benzodiazepine, derivatives |
| Fibrates | Medication | ATC:C10AB | Fibrates |
| Ezetimibe | Medication | ATC:C10AX09 | Ezetimibe |
| Aspirin | Medication | ATC:B01AC06 | Aspirin |
| ACEi/ARB | Medication | ATC:C09 | Agents acting on the renin-angiotensin system |
| β-Blockers | Medication | ATC:C07 | Beta blocking agents |
| NSAID | Medication | ATC: M01A | Anti-inflammatory agents, non-steroids |
| SGLT2 inhibitors | Medication | ATC: A10BK | Sodium-glucose co-transporter 2 (sglt2) inhibitors |
| GLP-1 RA | Medication | ATC: A10BJ | Glucagon-like peptide-1 (glp-1) analogues |
| BMI | Laboratory | TNX:9083 | BMI |
| Total cholesterol | Laboratory | TNX:9000 | Cholesterol [Mass/volume] in serum or plasma |
| LDL, cholesterol | Laboratory | TNX:9002 | Cholesterol in LDL [Mass/volume] in serum or plasma |
| HDL, cholesterol | Laboratory | TNX:9001 | Cholesterol in HDL [Mass/volume] in serum or plasma |
| Triglyceride | Laboratory | TNX:9004 | Triglyceride [Mass/volume] in serum, plasma or blood |

**Abbreviations**: ACEi, angiotensin-converting enzyme inhibitors; ARB, angiotensin receptor blockers; BMI, body mass index; CCB, calcium channel blockers; GLP-1 RA, glucagon-like peptide-1 receptor agonists; HDL, high-density lipoprotein; HMG-CoA, hydroxymethylglutaryl-coenzyme A; LDL, low-density lipoprotein; NSAID, nonsteroidal anti-inflammatory drugs; SGLT2, sodium-glucose cotransporter-2.

**Supplemental Table 4.** Diagnostic codes used in the definition of outcomes.

| **Outcomes** | **Category** | **Code** | **Description** |
| --- | --- | --- | --- |
| 1. **The primary outcome** | | | |
| Depression | Diagnosis | ICD10CM:F32.0 | Major depressive disorder, single episode, mild |
|  | Diagnosis | ICD10CM:F32.1 | Major depressive disorder, single episode, moderate |
|  | Diagnosis | ICD10CM:F32.2 | Major depressive disorder, single episode, severe without psychotic features |
|  | Diagnosis | ICD10CM:F32.3 | Major depressive disorder, single episode, severe with psychotic features |
|  | Diagnosis | ICD10CM:F32.8 | Other depressive episodes |
|  | Diagnosis | ICD10CM:F32.9 | Major depressive disorder, single episode, unspecified |
|  | Diagnosis | ICD10CM:F33.0 | Major depressive disorder, recurrent, mild |
|  | Diagnosis | ICD10CM:F33.1 | Major depressive disorder, recurrent, moderate |
|  | Diagnosis | ICD10CM:F33.2 | Major depressive disorder, recurrent severe without psychotic features |
|  | Diagnosis | ICD10CM:F33.3 | Major depressive disorder, recurrent, severe with psychotic symptoms |
|  | Diagnosis | ICD10CM:F34.1 | Dysthymic disorder |
|  | Diagnosis | ICD10CM:F43.21 | Adjustment disorder with depressed mood |
| 2. **All-cause mortality** | | | |
| All-cause mortality | Demographics | Deceased | Deceased |
|  | Diagnosis | ICD10CM:R99 | Ill-defined and unknown cause of mortality |
| 3. **Outcomes of specific depression subtypes** | | | |
| Major depressive disorder | Diagnosis | ICD10CM:F32.0 | Major depressive disorder, single episode, mild |
|  | Diagnosis | ICD10CM:F32.1 | Major depressive disorder, single episode, moderate |
|  | Diagnosis | ICD10CM:F32.2 | Major depressive disorder, single episode, severe without psychotic features |
|  | Diagnosis | ICD10CM:F32.3 | Major depressive disorder, single episode, severe with psychotic features |
|  | Diagnosis | ICD10CM:F32.9 | Major depressive disorder, single episode, unspecified |
|  | Diagnosis | ICD10CM:F33.0 | Major depressive disorder, recurrent, mild |
|  | Diagnosis | ICD10CM:F33.1 | Major depressive disorder, recurrent, moderate |
|  | Diagnosis | ICD10CM:F33.2 | Major depressive disorder, recurrent severe without psychotic features |
|  | Diagnosis | ICD10CM:F33.3 | Major depressive disorder, recurrent, severe with psychotic symptoms |
| Dysthymic disorder | Diagnosis | ICD10CM:F34.1 | Dysthymic disorder |
| Adjustment disorder | Diagnosis | ICD10CM:F43.21 | Adjustment disorder with depressed mood |
| 4. **Positive control outcome** | | | |
| Type 2 diabetes mellitus | Diagnosis | ICD10CM:E11 | Type 2 diabetes mellitus |
| 5. **Negative control outcomes** |  |  |  |
| Acute appendicitis | Diagnosis | ICD10CM:K35 | Acute appendicitis |
| Melanoma | Diagnosis | ICD10CM:C43 | Malignant melanoma of skin |
| Epistaxis | Diagnosis | ICD10CM:R04.0 | Epistaxis |

**Supplemental Figure 1.** Graphical illustration of the study design.

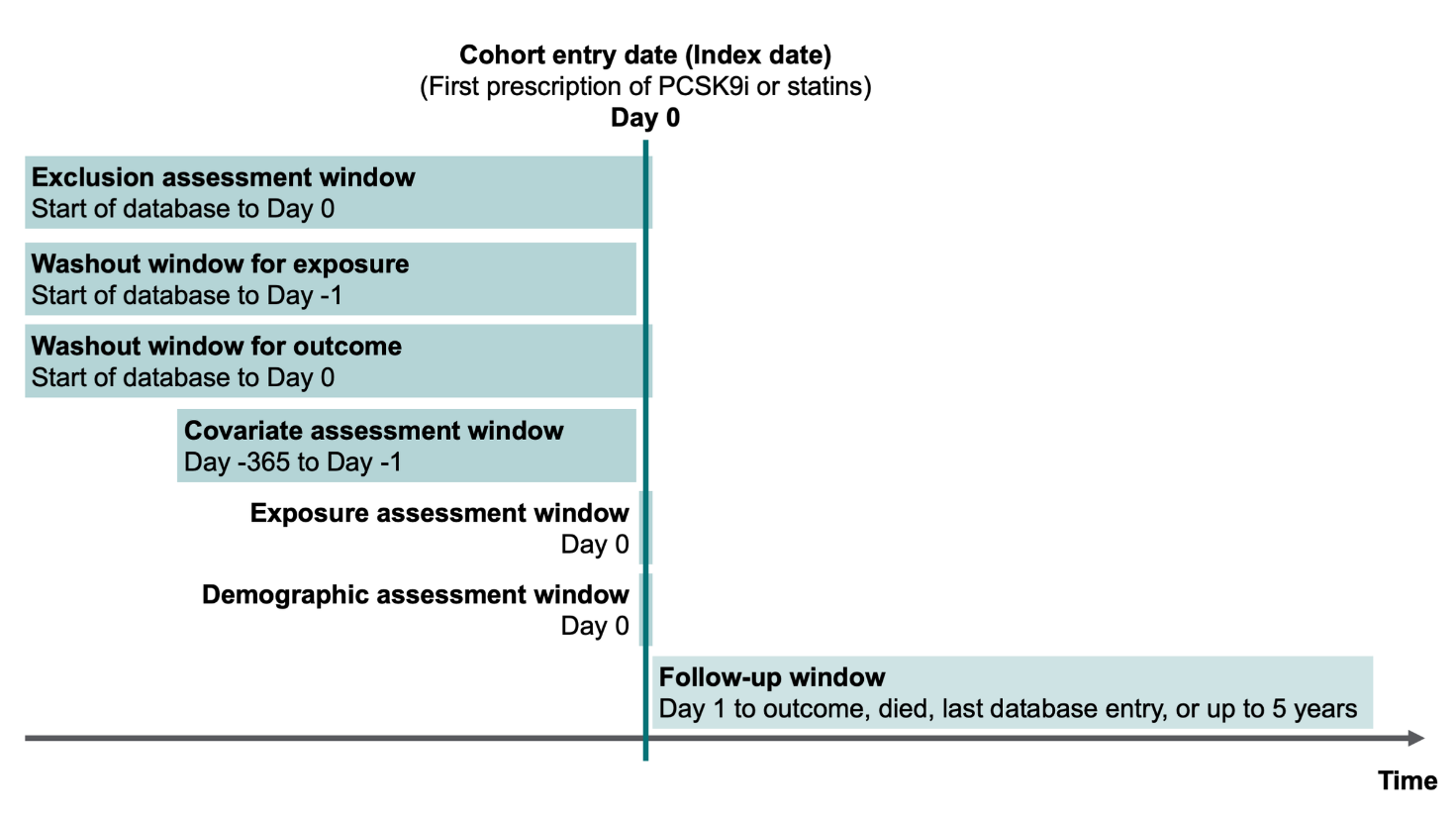

**Supplemental Figure 2.** Forest plot depicting hazard ratios for depression incidence in sensitivity and validation analyses.

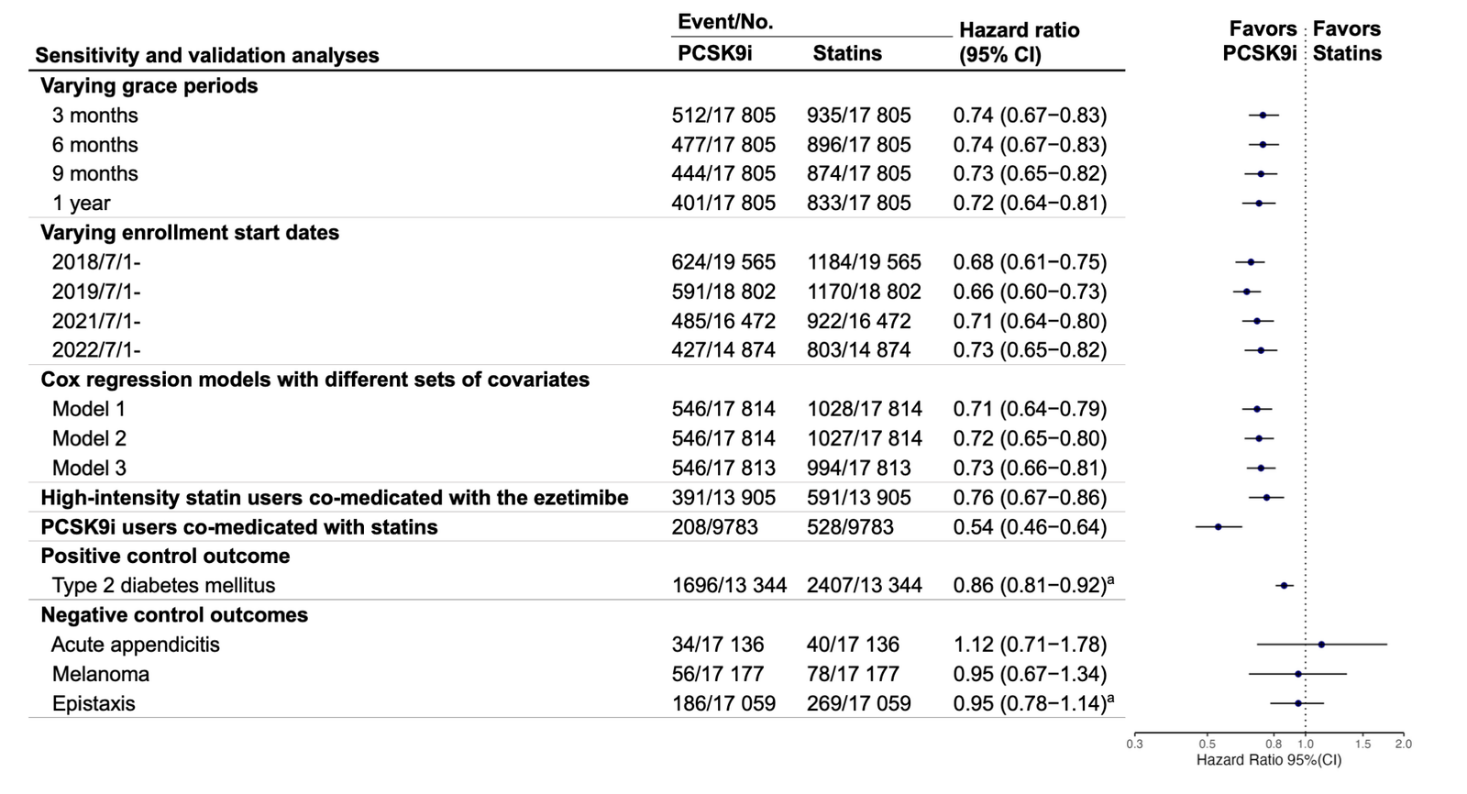

Grace periods are amounts of time (3, 6, 9, or 12 months) calculated from the individual's index date, after which follow-up time was considered to begin. Model 1: propensity score matching was performed on age at index, sex, race. Model 2: propensity score matching was performed on age at index, sex, race, social economic status, lifestyles, medical utilization. Model 3: propensity score matching was performed on age at index, sex, race, social economic status, lifestyles, medical utilization, comorbidities and medications use. High-intensity statin users co-medicated with ezetimibe were defined as patients with hyperlipidemia who were prescribed ezetimibe on or within 1 month of the index date of high-intensity statin (atorvastatin 40–80 mg, or rosuvastatin 20–40 mg) use. PCSK9i users co-medicated with statins were defined as patients with hyperlipidemia prescribed statins on or within 1 month of the index date of PCSK9i.

^a^proportionality < .05.

**Abbreviations**: CI, confidence interval; HR, hazard ratio; PCSK9i, proprotein convertase subtilisin/kexin type 9 inhibitors.

**STROBE Statement**—Checklist of items that should be included in reports of ***cohort studies***

|  | Item No | Recommendation | Page |
| --- | --- | --- | --- |
| **Title and abstract** | 1 | (*a*) Indicate the study’s design with a commonly used term in the title or the abstract | 1-3 |
|  |  | (*b*) Provide in the abstract an informative and balanced summary of what was done and what was found |  |
| Introduction | | |  |
| Background/rationale | 2 | Explain the scientific background and rationale for the investigation being reported | 4-6 |
| Objectives | 3 | State specific objectives, including any prespecified hypotheses | 5-6 |
| Methods | | |  |
| Study design | 4 | Present key elements of study design early in the paper | 16-21 |
| Setting | 5 | Describe the setting, locations, and relevant dates, including periods of recruitment, exposure, follow-up, and data collection | 16-17 |
| Participants | 6 | (*a*) Give the eligibility criteria, and the sources and methods of selection of participants. Describe methods of follow-up | 17-18 |
|  |  | (*b*) For matched studies, give matching criteria and number of exposed and unexposed |  |
| Variables | 7 | Clearly define all outcomes, exposures, predictors, potential confounders, and effect modifiers. Give diagnostic criteria, if applicable | 18 |
| Data sources/ measurement | 8* | For each variable of interest, give sources of data and details of methods of assessment (measurement). Describe comparability of assessment methods if there is more than one group | 19-21 |
| Bias | 9 | Describe any efforts to address potential sources of bias | 19-21 |
| Study size | 10 | Explain how the study size was arrived at | 16 |
| Quantitative variables | 11 | Explain how quantitative variables were handled in the analyses. If applicable, describe which groupings were chosen and why | 19-21 |
| Statistical methods | 12 | (*a*) Describe all statistical methods, including those used to control for confounding | 19-21 |
|  |  | (*b*) Describe any methods used to examine subgroups and interactions |  |
|  |  | (*c*) Explain how missing data were addressed |  |
|  |  | (*d*) If applicable, explain how loss to follow-up was addressed |  |
|  |  | (*e*) Describe any sensitivity analyses |  |
| Results | | |  |
| Participants | 13* | (a) Report numbers of individuals at each stage of study—eg numbers potentially eligible, examined for eligibility, confirmed eligible, included in the study, completing follow-up, and analysed | 7 |
|  |  | (b) Give reasons for non-participation at each stage |  |
|  |  | (c) Consider use of a flow diagram |  |
| Descriptive data | 14* | (a) Give characteristics of study participants (eg demographic, clinical, social) and information on exposures and potential confounders | 7 |
|  |  | (b) Indicate number of participants with missing data for each variable of interest |  |
|  |  | (c) Summarise follow-up time (eg, average and total amount) |  |
| Outcome data | 15* | Report numbers of outcome events or summary measures over time | 7-8 |
| Main results | 16 | (*a*) Give unadjusted estimates and, if applicable, confounder-adjusted estimates and their precision (eg, 95% confidence interval). Make clear which confounders were adjusted for and why they were included | 7-8 |
|  |  | (*b*) Report category boundaries when continuous variables were categorized |  |
|  |  | (*c*) If relevant, consider translating estimates of relative risk into absolute risk for a meaningful time period |  |
| Other analyses | 17 | Report other analyses done—eg analyses of subgroups and interactions, and sensitivity analyses | 8-10 |
| Discussion | | |  |
| Key results | 18 | Summarise key results with reference to study objectives | 11 |
| Limitations | 19 | Discuss limitations of the study, taking into account sources of potential bias or imprecision. Discuss both direction and magnitude of any potential bias | 13-15 |
| Interpretation | 20 | Give a cautious overall interpretation of results considering objectives, limitations, multiplicity of analyses, results from similar studies, and other relevant evidence | 11-15 |
| Generalisability | 21 | Discuss the generalisability (external validity) of the study results | 13-15 |
| Other information | | |  |
| Funding | 22 | Give the source of funding and the role of the funders for the present study and, if applicable, for the original study on which the present article is based | 27 |

*Give information separately for exposed and unexposed groups.

**Note:** An Explanation and Elaboration article discusses each checklist item and gives methodological background and published examples of transparent reporting. The STROBE checklist is best used in conjunction with this article (freely available on the Web sites of PLoS Medicine at http://www.plosmedicine.org/, Annals of Internal Medicine at http://www.annals.org/, and Epidemiology at http://www.epidem.com/). Information on the STROBE Initiative is available at http://www.strobe-statement.org.

**TARGET Checklist of Recommended Items to Address in Reports of Studies Emulating a Target Trial^a^**

| **Item No** | **Checklist item** |  | | | **Page** |
| --- | --- | --- | --- | --- | --- |
| **Abstract** | | | | | |
| 1 | a | Identify that the study attempts to emulate a target trial using observational data. State the study objectives and briefly summarize the specified target trial. | | | 3 |
|  | b | Report the data sources used for emulation. | | | 3 |
|  | c | Summarize key assumptions, statistical methods, findings and conclusions. | | | 3 |
| **Introduction** | | | | | |
| 2 | Background | Describe the scientific background of the study and the gap in knowledge. | | | 4-6 |
| 3 | Causal question | Summarize the causal question. | | | 5-6 |
| 4 | Rationale | Describe the rationale for emulating a target trial with the available data. Cite randomized trials informing the design of the target trial if applicable. | | | 5-6 |
| **Methods** | | | | | |
| 5 | Data source | Cite the data sources contributing to the analyses and for each one describe the following: original purpose, type, the geographical locations, setting, and time period. If relevant, describe how the data were linked or pooled. | | | 16 |
| **Target trial specification (item 6) and target trial emulation (item 7)** | | | | | |
| 6 | Specify the components of the target trial protocol that would answer the causal question. | | 7 | Describe how the components of the target trial protocol were emulated with the observational data, including how all variables were measured or ascertained. | 17-18  eTable 1 |
| Eligibility criteria | | | | | |
| 6a | Describe the eligibility criteria. | | 7a | Describe how the eligibility criteria were operationalized with the data. | 17-18  eTable 1 |
| Treatment strategies | | | | | |
| 6b | Describe the treatment strategies that would be compared. | | 7b | Describe how the treatment strategies were operationalized with the data. | 17-18  eTable 1 |
| Assignment procedures | | | | | |
| 6c | Report that eligible individuals would be randomly assigned to treatment strategies and may be aware of their treatment allocation. | | 7c | Describe how assignment to treatment strategies was operationalized with the data. | 17-18  eTable 1 |
| Follow-up | | | | | |
| 6d | Clarify that follow-up would start at time of assignment to the treatment strategies. Specify when follow-up would end. | | 7d | Clarify that follow-up starts at the time individuals were assigned to the treatment strategies. Describe how the end of follow-up was operationalized with the data. | 17-18  eTable 1 |
| Outcomes | | | | | |
| 6e | Describe the outcomes | | 7e | Describe how the outcomes were operationalized with the data. | 19  eTable 1 |
| Causal contrasts | | | | | |
| 6f | Describe the causal contrasts of interest, including effect measures. | | 7f | Describe how the causal contrasts were operationalized with the data, including effect measures. | eTable 1 |
| Identifying assumptions | | | | | |
| 6g | Describe assumptions that would be made to identify each causal estimand. Describe the variables, if any, related to these assumptions. | | 7g.i | For each causal estimand, describe assumptions made to identify it, including assumptions regarding baseline confounding due to lack of randomization. | eTable 1 |
|  |  | | 7g.ii | Describe how the variables related to these assumptions were operationalized with the data. | eTable 1 |
| Data analysis plan | | | | | |
| 6h | For each causal estimand, describe the data analysis procedures and any associated statistical modeling assumptions, including approaches for handling missing data. | | 7h.i | For each causal estimand, describe the data analysis procedures and any associated statistical modeling assumptions, including approaches for handling missing data. | 19-21  eTable 1 |
|  |  | | 7h.ii | For each causal estimand, describe any additional analyses conducted to assess the sensitivity of the results to the choice of operationalizations, assumptions and analysis. | 19-21  eTable 1 |
| **Results** | | | | | |
| 8 | Participant selection | Report numbers of individuals assessed for eligibility, eligible, and assigned to each treatment strategy. A flow diagram is strongly recommended. | | | 7,  Figure 1 |
| 9 | Baseline data | Describe the distribution of characteristics of individuals at baseline, by treatment strategy. | | | 7,  Table 1 |
| 10 | Follow-up | Summarize length of follow-up and describe reasons for end of follow-up for each treatment strategy and causal contrast. | | | 7 |
| 11 | Missing data | Describe the frequency of missing data in all variables, by treatment strategy when applicable. | | | 7,  Table 1 |
| 12 | Outcomes | Describe the frequency or distribution of each outcome, by treatment strategy. | | | 7-8 |
| 13 | Effect estimates | Report the effect estimates for each causal contrast with corresponding measures of precision, including both absolute and relative measures of effect, when applicable. | | | 7-8, Table 2 |
| 14 | Additional analyses | Report results of all analyses to assess the sensitivity of the estimates to choices in operationalizations, assumptions and analysis. | | | 8-10 |
| **Discussion** | | | | | |
| 15 | Interpretation | Provide an interpretation of the key findings. | | | 11 |
| 16 | Limitations | Discuss the limitations of the study considering differences between the target trial and its emulation and the plausibility of assumptions, including assumptions regarding baseline confounding due to lack of randomization. | | | 13 |
| **Other information** | | | | | |
| 17 | Ethics | Provide the institutional research board or ethics committee that approved the study and approval numbers, if relevant. | | | 17 |
| 18 | Registration | State whether, when, and where the study protocol was registered. | | | NA |
| 19 | Sharing of study materials | Provide information on whether data, analytic code and/or other materials are accessible, and where and how they can be accessed. | | | NA |
| 20 | Funding sources | Provide the sources of funding and detail the role of the funders in the design, conduct and reporting of the study. | | | 27 |
| 21 | Conflicts of interest | State any conflicts of interest and financial disclosures for all authors. | | | 28 |

**^a^**Republished with permission from the TARGET group. The TARGET Checklist is licensed by the TARGET group under the Creative Commons Attribution-NoDerivs (CC BY-ND) 4.0 International license.
